# A statistical immune correlates of protection model for predicting efficacy from neutralizing antibody titers to establish immunobridging of monoclonal antibodies for prevention of COVID-19

**DOI:** 10.1101/2025.07.24.25332145

**Authors:** Ilker Yalcin, Leijun Hu, Brandyn West, Kristin Narayan, Anna Holmes, Mark A. Wingertzahn

## Abstract

**Background:** Neutralizing antibody titers are recognized as an acceptable surrogate efficacy endpoint for immunobridging next-generation monoclonal antibodies (mAbs) to those with demonstrated clinical efficacy for the prevention of COVID-19. However, titers measured at early time points after dosing overestimate the titer levels required for clinical protection, but long-term efficacy data is limited due to continued evolution of SARS-CoV-2 and loss of activity of previously effective mAbs against emerging variants. To address these challenges, we set out to develop a predictive tool for efficacy using neutralizing titers to establish immunobridging for mAbs for prevention of COVID-19.

**Methods:** Drug concentration and clinical efficacy data collected over 12 months following administration of pemivibart in the phase 3 CANOPY trial for prevention of COVID-19 were used to develop a Cox proportional hazards model with time-varying covariate as a statistical immune correlates of protection (CoP) model. The time-varying covariate was estimated serum virus neutralizing antibody (sVNA) activity, which changes over time in response to the emergence and prevalence of different SARS-CoV-2 variants. sVNA was estimated at 2-week intervals by integrating drug concentration data with weighted average IC_50_ (half-maximal inhibitory concentration) values of the circulating variant population. This model was used to predict clinical efficacy in both immunocompromised (IC) and non-immunocompromised (non-IC) populations based on sVNA titers.

**Results:** Efficacy increased with antibody titer in a non-linear manner, with smaller incremental gains at higher concentrations. Predicted efficacy was lower at all titer levels in the IC cohort. Model-derived estimates aligned well with observed infection outcomes and external analyses, supporting model validity. Based on the model, a sVNA titer of 1:500 predicted an estimated 50% efficacy in IC and 70% efficacy in non-IC populations, where efficacy is defined as the relative reduction in the risk of COVID-19 infection.

**Conclusion:** The Cox model provides a foundation for evaluation of suitable neutralizing titer targets to guide dosing, predict estimated clinical benefit, and support immunobridging in both IC and non-IC populations.

## Introduction

The use of immune correlates of protection (CoP) to infer vaccine efficacy is a scientifically validated and regulatory-endorsed strategy in vaccine approvals. A well-established precedent comes from influenza vaccine development, where a hemagglutination inhibition (HI) titer of 1:40 has historically been accepted as conferring approximately 50% protection against symptomatic infection (Hobson 1972; Coudeville 2010). This threshold has informed the evaluation and approval of influenza vaccines. Once a CoP is agreed upon by regulatory authorities, it enables the demonstration of efficacy for new vaccines through traditional non-inferiority criteria. This approach is referred to as immunobridging and it involves comparing either the proportion of vaccine recipients who achieve the protective threshold or the geometric mean titers (GMT) relative to those elicited by a licensed comparator vaccine.

More recently, similar principles have been applied to COVID-19. Emerging data demonstrate a robust quantitative relationship between neutralizing antibody levels and observed clinical efficacy across multiple vaccine platforms (Khoury 2021; Schmidt 2023). These findings support the use of immunological markers such as neutralizing titers as reliable surrogate endpoints for vaccine protection. In the setting of rapidly evolving SARS-CoV-2 variants and the global shift away from placebo-controlled trials, the immunobridging approach offers a scientifically valid and ethically appropriate pathway for vaccine assessment and regulatory decision-making.

CoPs are not limited to active immunization such as vaccines; they are also relevant to passive immunization strategies, including monoclonal antibodies. Serum virus neutralizing antibody (sVNA) titers and serum drug concentrations serve as reliable surrogate endpoints to support efficacy across mAb platforms. In the absence of placebo-controlled trials, especially in immunocompromised (IC) populations or against ever-emergent variants, these markers enable variant-specific or population-based immunobridging, aligned with regulatory precedent and model-informed drug development strategies. The relationship between sVNA titers derived from monoclonal antibodies and protection against COVID-19 has been explored using clinical study data in immune naïve individuals prior to widespread vaccination and immune exposure (Schmidt 2023; Stadler 2023).

Pemivibart was the first (and to date, only) SARS-CoV-2 mAb to effectively use the immunobridging approach to obtain EUA for prevention of COVID-19 (Schmidt 2024; Invivyd 2025). Data from a single-arm cohort (Cohort A) of the 2-cohort CANOPY phase 3 trial (NCT06039449) including 306 IC individuals were used to establish immunobridging. Cohort B of the study randomized 482 non-IC individuals in a 2:1 ratio to receive either pemivibart or placebo. In addition to assessing safety, the trial evaluated the efficacy of pemivibart in preventing symptomatic COVID-19. Both cohorts were conducted concurrently, and all participants were followed for 12 months.

Participants received 4500 mg of pemivibart intravenously on Day 1 and Month 3. The analysis of the primary endpoint in Cohort A compared sVNA titers calculated for pemivibart (pharmacokinetic [PK] concentration / IC50 of select relevant variant) with those calculated for the prototype parent mAb adintrevimab against the Delta variant predominant during the EVADE trial.

In the EVADE study (NCT04859517), adintrevimab 300 mg provided 71% protection from symptomatic COVID-19 through 3 months (Ison 2023). EVADE was prematurely suspended due to the circulating variant shift from Delta to Omicron lineages, in which adintrevimab demonstrated a decline in neutralizing activity. The titer value of adintrevimab against Delta at Month 3 (1:3514) was used as the immunobridging target, as substantial data from later time points were unavailable.

Immunobridging in the CANOPY IC Cohort A was achieved if the lower limit of the 90% confidence interval of the geometric mean ratio (GMR) between the selected titer target and the mAb/variant-specific sVNA titer was >0.80. Strict immunobridging of pemivibart against the JN.1 variant (dominant at the time of analysis) to adintrevimab through 3 months was not met (GMR = 0.70 [90% CI: 0.68–0.72]); however, a complementary immunobridging method using data from a meta-analysis showed that the titer values for pemivibart were consistent with the ranges observed in previous clinical trials of other mAbs with proven clinical efficacy (Follmann 2023; Schmidt 2023; Stadler 2023; Schmidt 2024). Authorization was granted based on the totality of evidence (Schmidt 2024).

Although strict immunobridging predicting pemivibart efficacy at 3 months was not met, pemivibart exhibited a 93.7% relative risk reduction (RRR) in prevention of RT-PCR-confirmed COVID-19 at 3 months and a 74.1% RRR over 12 months against a range of dominant circulating variants compared with placebo in the CANOPY non-IC Cohort B (Table 1). The efficacy results indicate that the immunobridging target was set too high to accurately predict clinically meaningful efficacy. Notably, the clinical efficacy of pemivibart through Month 12 was associated with neutralizing titer levels as low as approximately 1:50 against KP.3.1.1 (Table 1), the dominant circulating variant at that time in the study, which is substantially lower than the previously considered target of 1:3514 for meaningful efficacy (Schmidt 2024). These data demonstrate the importance of collecting correlative data at later timepoints when serum concentrations have waned to fully characterize the relationship between sVNA titers and clinical efficacy, which was not possible in the EVADE study due to limited follow-up.

**Table 1:** Efficacy and sVNA Titers of Pemivibart Over a 12-month Period in the CANOPY Study (Cohort B)

| Period (months) | Placebo |  |  | Pemivibart |  |  | sRRR (%) | p-value | Geometric Mean Pemivibart Concentration (ug/mL) | Predominant Variant at Period End | sVNA at Period End |
| --- | --- | --- | --- | --- | --- | --- | --- | --- | --- | --- | --- |
|  | n | COVID-19 Cases<sup>a</sup> | % | n | COVID-19 Cases<sup>a</sup> | % |  |  |  |  |  |
| 0 - 3 | 158 | 8 | 5.1 | 315 | 1 | 0.3 | 93.7 | 0.0086 | 172 | HV.1 | 4310 |
| 0 - 6 | 158 | 19 | 12.0 | 315 | 6 | 1.9 | 84.2 | <0.0001 | 209 | JN.1 | 2822 |
| 0 - 9 | 158 | 20 | 12.7 | 315 | 9<sup>b</sup> | 2.9 | 77.2 | <0.0001 | 48 | KP.3 | 216 |
| 0 - 12 | 158 | 29 | 18.4 | 315 | 15<sup>b</sup> | 4.8 | 74.1 | <0.0001 | 11 | KP.3.1.1 | 47 |

sRRR = standardized relative risk reduction; sVNA = serum virus neutralizing antibody
<sup>a</sup> COVID-19 cases defined as RT-PCR-confirmed symptomatic COVID-19, COVID-19-related hospitalization, or all-cause death
<sup>b</sup> Includes one COVID-19-unrelated death

Data from other studies have likewise supported efficacy with lower neutralizing titers. In an exploratory analysis of the post-Omicron portion of EVADE, adintrevimab provided a 60% RRR against Omicron BA.1 at 1 month; the associated calculated titer was 1:33 (Schmidt 2023). This target is also in the range of what was seen with tixagevimab/cilgavimab 300 mg/300 mg against Omicron BA.1, where titers of 1:71 at 6 months were expected to provide slightly greater than 70% protection against symptomatic disease (Dejnirattisai 2022; AstraZeneca 2023).

Cumulative data with SARS-CoV-2-directed mAbs indicate that calculated titers can serve as surrogate immunobridging endpoints that are highly likely to predict efficacy; however, the immunobridging target must be refined to more precisely reflect the titers that likely result in clinical protection based on neutralizing titers and clinical efficacy analysis over a continuum of timepoints.

Most clinical efficacy data to date comes from studies performed in non-IC populations. The only randomized phase 3 trial evaluating the efficacy of a mAb in IC individuals is the SUPERNOVA trial that assessed sipavibart 300 mg for the prevention of symptomatic COVID-19 (Haidar 2025). The SUPERNOVA trial demonstrated a RRR of 42.9% (p=0.0012) of sipavibart 300 mg in preventing symptomatic COVID-19 in IC participants by susceptible Omicron XBB- and JN.1-related variants (Haidar 2025). Efficacy was modest compared to mAb studies in non-IC participants (CANOPY, EVADE, or PROVENT), where RRRs of >70% were observed. These data may suggest that the overall level of efficacy achieved in IC individuals may be lower at similar neutralizing titer levels.

The methodology described below proposes a predictive tool for efficacy using neutralizing titers to establish immunobridging for mAbs developed on the same structural platform for prevention of COVID-19 in both IC and non-IC populations.

## Methods

Data from both cohorts of the CANOPY trial were used to develop a Cox proportional hazards model with time-varying covariate as a statistical immune CoP model. The time-varying covariate was calculated sVNA, which changes over time in response to the emergence and prevalence of different SARS-CoV-2 variants. sVNA was calculated at 2-week intervals by integrating drug concentration data with weighted IC_50_ values of the circulating variants as detailed below.

Two separate regression models were constructed for IC and non-IC participants using clinical data up to 12 months and estimated pemivibart concentrations from Cohort A (IC) and Cohort B (non-IC) participants, respectively. The models used time from the first dosing to the first RT-PCR-confirmed symptomatic COVID-19 as the dependent variable and daily weighted titer levels as the independent variable. The predictor (independent) variable was calculated sVNA levels estimated from the drug concentrations divided by pseudovirus neutralization assay IC_50_ against a relevant variant.

As Cohort A did not have a placebo control, to enable a comparable estimation of clinical efficacy, placebo data from Cohort B were used as the control group in the model. This is the limitation of this model for IC participants. However, the fact that both CANOPY cohorts were conducted concurrently helps mitigate this limitation by providing a temporally aligned reference from the placebo arm in Cohort B. In addition, the partially contemporaneous placebo-controlled SUPERNOVA trial, which evaluated sipavibart in an IC population during the JN.1 surge, reported a similar 6-month attack rate in its comparator arm (12% in CANOPY vs 11% in SUPERNOVA)(Haidar 2025), further supporting the external validity of the CANOPY Cohort B placebo data as a reference for modeling efficacy in the IC population.

### Concept of weighted IC_50_

Clinical studies span periods with multiple circulating variants. Figure 1 presents the SARS-CoV-2 variants during the CANOPY study time frame. Five predominant SARS-CoV-2 variants circulated during the study along with numerous other variants with notably different IC_50_ values. A neutralizing titer variable was identified and included in the Cox model for calculating neutralizing titer to account for the complexity introduced by variant variability. We developed the weighted average IC_50_ concentration for a given timeframe by summing the product of each variant’s proportion (w) and its closest representative pseudovirus IC_50_ value over the timeframe:

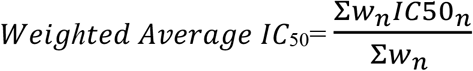

**Figure 1:**
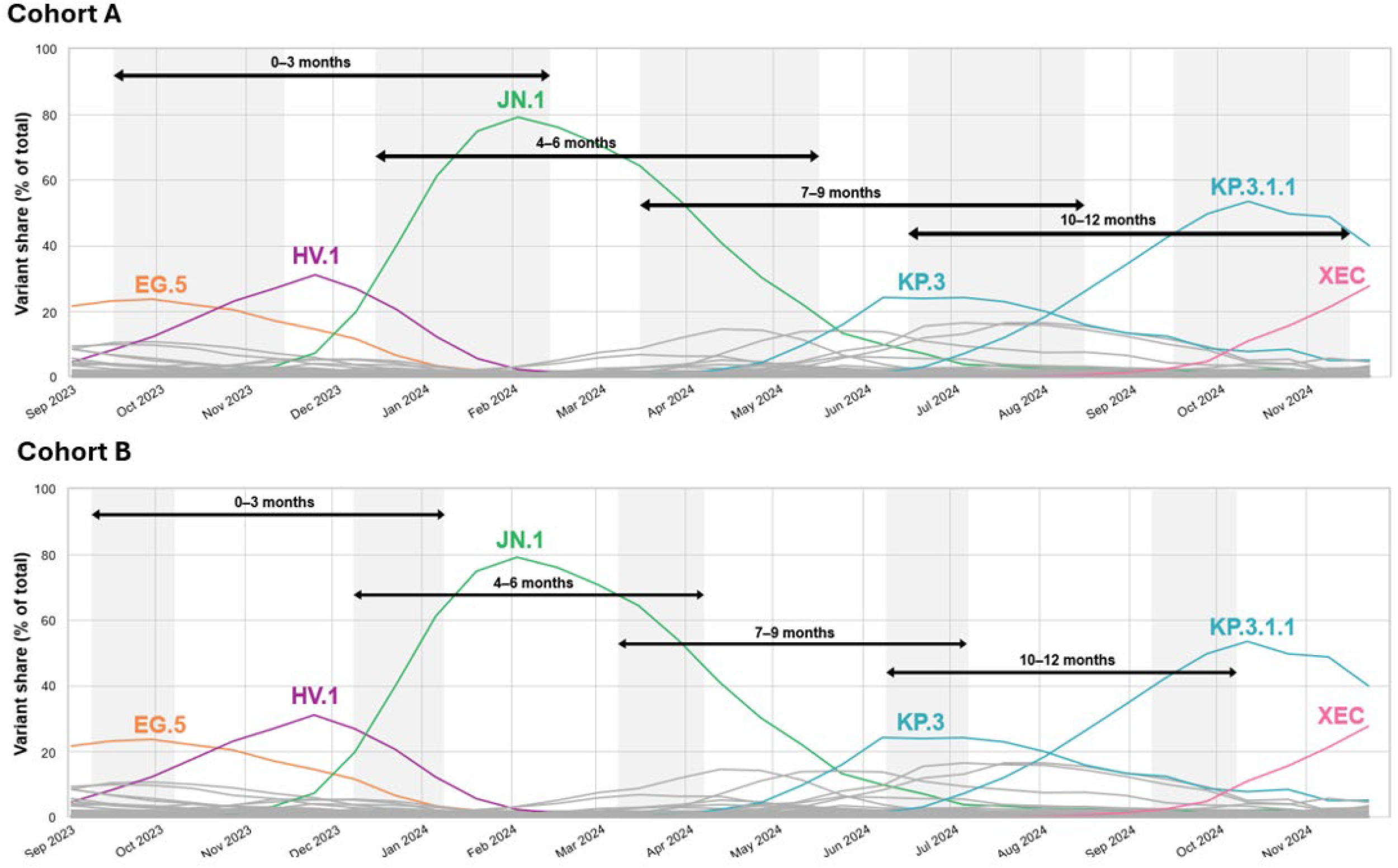
SARS-CoV-2 Predominant Variants During CANOPY Study Time Frame. CDC=Centers for Disease Control and Prevention. Note: Shaded areas represent the time required to complete the enrollment. Data for circulating proportions of CDC-tracked variants accessed at https://data.cdc.gov/Laboratory-Surveillance/SARS-CoV-2-Variant-Proportions/jr58-6ysp/about_data on 24-Jan-25. Source: Internal data on file and (CDC 2025).

Variant proportions were sourced from the CDC variant tracker; pemivibart’s pseudovirus IC_50_ values were determined by Monogram Biosciences (Invivyd 2025). When the IC_50_ was unavailable for a variant, it was assigned the closest match based on phylogenetic proximity using pangolin lineage analysis. Variants with minor fractions classified as “other” by the CDC were excluded. Given pemivibart’s robust IC_50_ dataset covering major variants, this method provides a reasonable approximation.

### Log-transformed daily weighted neutralizing titer as independent variable

Daily weighted neutralizing titer was obtained by dividing average pemivibart serum concentration of each day by the weighted IC_50_. The average pemivibart concentration of each day from the first dose was estimated using post-hoc PK parameters of a population PK model developed for CANOPY participants; weighted IC_50_ value was updated every 2 weeks (the same frequency by which the CDC variant tracker updates) based on calendar days through the study period. For each participant, the predicted daily average pemivibart concentration and weighted IC_50_ value were matched by calendar days and the correspondent daily weighted neutralizing titer was calculated. sVNA titers were further log-transformed (log_10_).

### Time-varying Cox proportional hazards model

The daily risk of acquiring COVID-19 from the first dose through 365 days was estimated using a time-varying Cox proportional hazards model, the hazard function is specified as:

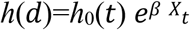

where *h*_*0*_*(t)* is the baseline hazard, *t* is the number of days after the first dose, and *X*_*t*_ is the log_10_ weighted sVNA on day *t*. Clinical Efficacy (CE) as a function of weighted titer (*X*) is specified as:

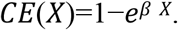

Since the weighted titer is derived from the predicted daily pemivibart PK concentration, placebo participants were assigned to a *log*_*10*_ calculated titer of 0. For the model developed using Cohort B data, this zero-value assignment effectively created a natural dichotomous treatment variable. As a result, the model yielded an interpretable estimate for 1 minus the hazard ratio (1 – HR), representing the relative reduction in the risk of symptomatic COVID-19 associated with pemivibart exposure. Model fitting was conducted using R version 4.3.3 (R Core Team 2024).

This model is both Invivyd platform mAb- and variant-agnostic, allowing it to be applied to any variant with a known IC_50_.

## Results

The fitted model using the data in CANOPY Study Cohort B yielded *β* = - 0.4384 (standard error = 0.1009, likelihood ratio test p-value < 0.0001). The equation for predicting clinical efficacy from the titer level becomes:

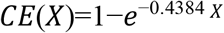

The model with Cohort A data combined with Cohort B placebo data yielded *β* = -0.2566 (standard error = 0.0832, likelihood ratio test p-value = 0.00172). The equation for predicting clinical efficacy from the titer level becomes:

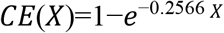

Figure 2 displays the predicted curves for weighted titers for Cohorts A and B, accompanied by percentile-based 95% confidence band derived from 10,000 bootstrap samples. Table 2 summarizes the efficacy predictions from the Cox model for selected target titers, indicating that a titer level of 1:50 corresponds to an estimated 52% efficacy in non-IC. In the absence of informative censoring, the clinical efficacy obtained from the Cox model (1 – HR) is expected to be similar to RRR. Table 2 also presents the number needed to treat (NNT) for 6-month protection, calculated as 1/(efficacy × 0.12) where 0.12 is the 6-month attack rate for placebo group.

**Table 2:** Predicted Efficacy for Selected sVNA Titer Targets Based on Time-varying Proportional Hazards Cox Model.

| Titer | Non-IC Predicted Efficacy (NNT) | IC Predicted Efficacy (NNT) |
| --- | --- | --- |
| 1:1000 | 73% (12) | 54% (16) |
| 1:500 | 69% (13) | 50% (17) |
| 1:400 | 68% (13) | 49% (17) |
| 1:300 | 66% (13) | 47% (18) |
| 1:200 | 64% (13) | 45% (19) |
| 1:100 | 58% (15) | 40% (21) |
| 1:50 | 52% (16) | 35% (24) |

sVNA = serum virus neutralizing activity; IC=immunocompromised; NNT = number needed to treat for 6-month protection = $1/(\text{Efficacy} \times 0.12)$

**Figure 2:**
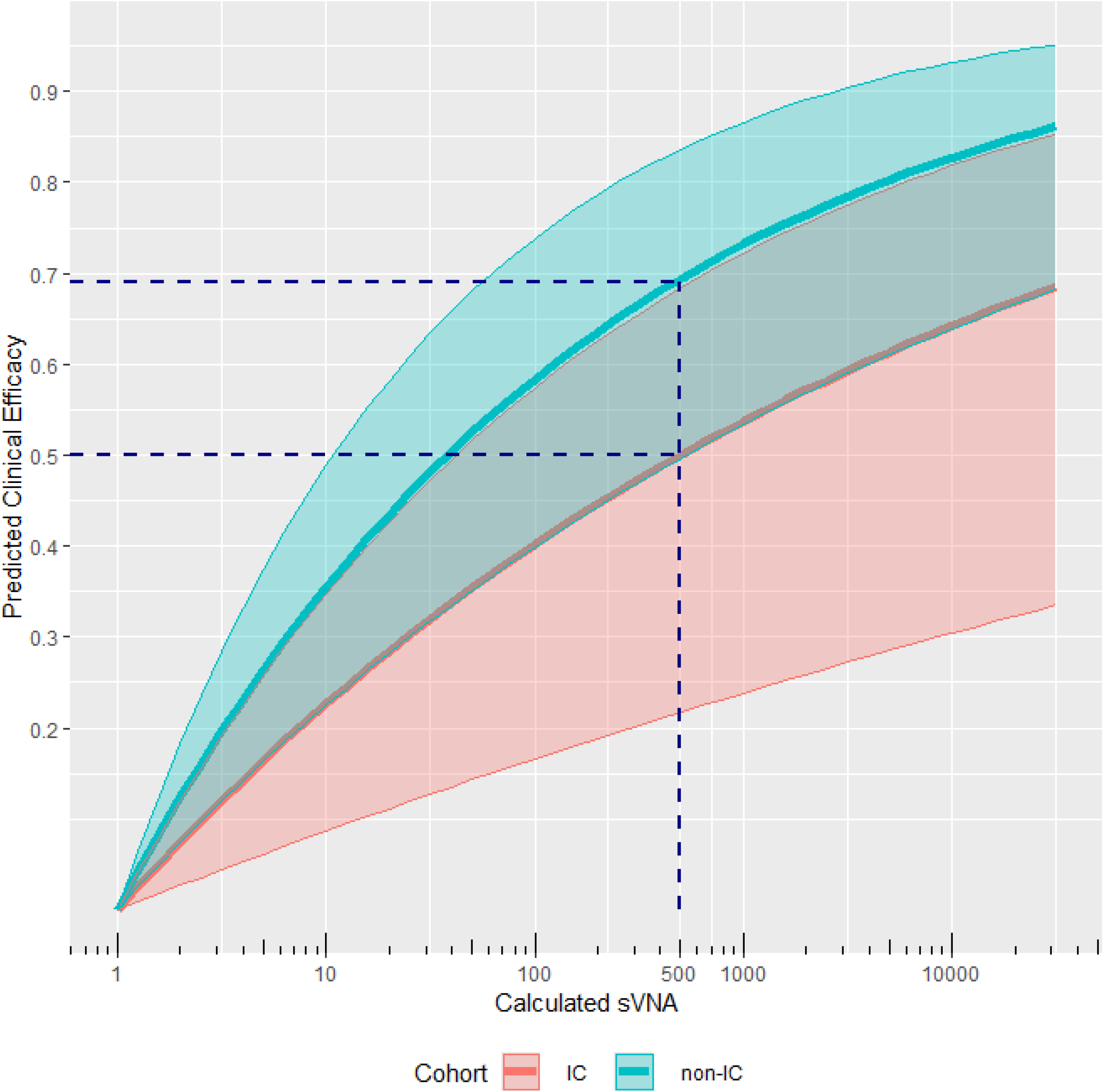
Time-varying Proportional Hazards Cox Model Efficacy Prediction Curves for Prevention of COVID-19. Efficacy and titer data from CANOPY Cohorts A (IC) and B (non-IC) were applied to a time-varying proportional hazards Cox model to generate an efficacy prediction curve. Blue dashed lines highlight the estimated efficacy for IC and non-IC populations at an sVNA level of 500.

Efficacy increased with sVNA titer in a non-linear manner, with smaller incremental gains at higher concentrations. Predicted efficacy was lower at all titer levels in the IC cohort. Model-derived estimates aligned well with observed infection outcomes and external analyses, supporting model validity.

In immunocompetent populations, maintaining target sVNA titers of 1:500 at the end of the dosing interval is associated with approximately 70% or greater clinical efficacy against currently dominant SARS-CoV-2 variants. This threshold provides a margin for continued protection against emerging variants with reduced susceptibility. For instance, a two-fold reduction in titer (to 1:250) results in less than a 5% decrease in predicted efficacy.

For IC populations, higher sVNA titers may be required to achieve protection levels comparable to those observed in immunocompetent individuals. At the end of the dosing interval, target titers of 1:500 are associated with approximately 50% or greater clinical efficacy against currently circulating variants. To achieve efficacy levels closer to those in immunocompetent populations, increased dosing frequency or higher dose density would likely be necessary to elevate titers accordingly.

## Discussion

In the phase 3 CANOPY trial, participants received 4500 mg of pemivibart intravenously on Day 1 and Month 3, with surrogate efficacy based on sVNA titers validating immunobridging as a reliable predictor of mAb effectiveness (Schmidt 2024; Wolfe 2025). However, the very high titer from the EVADE study that pemivibart was required to reach for successful immunobridging forced a very high dose of pemivibart to be administered. The clinical efficacy of pemivibart in preventing RT-PCR-confirmed COVID-19 over 12 months in placebo-controlled Cohort B suggested much lower titers are associated with clinical efficacy (Table 1).

By developing a time-varying Cox proportional hazards model based on CANOPY clinical data up to 12 months and estimated pemivibart concentrations from CANOPY Cohorts A and B participants, respectively, selection of dosage level and route based on modelling is possible for future Invivyd platform mAbs to optimize the benefit-risk profile.

Neutralizing titers as a surrogate marker have a clear mechanistic link to therapeutic effect, correlate with clinical outcomes, and are adaptable to the changing sublineages of COVID-19. The major merits of such an immunobridging target (Table 2) and associated titer-efficacy curve (Figure 2) are that: 1) The proposed target provides sufficient efficacy whereby real clinical benefit can still be conferred at lower titer levels that may result from drift in product IC_50_ over time, and 2) Since it is certain that the product IC_50_ estimates will change periodically with new variants, leveraging such a continuous quantitative relationship between IC_50_ and expected clinical efficacy provides an intelligible and actionable alteration to product description for healthcare providers, regulators, and sponsors to assess product risk/benefit in day-to-day clinical practice.

The predicted efficacy curve from the CANOPY Cohort B data is consistent with the findings from the logistic regression modelling introduced by Stadler et al. (Stadler 2023), who analyzed publicly available summary level data from randomized controlled trials evaluating mAbs for the prevention of symptomatic SARS-CoV-2 infection primarily in immunocompetent populations. By modeling the dose-response relationship using in vivo mAb concentrations (normalized by in vitro IC_50_ value) and corresponding protection levels over time, they showed that high neutralizing titers (e.g., >1:500, which corresponds to approximately 70% efficacy, compared to 69% efficacy in our Cox model) are consistently linked to strong prophylactic efficacy across multiple mAbs. Figure 3 presents the curve originally introduced by Stadler (Stadler 2023), updated with pemivibart data (Schmidt 2024).

**Figure 3:**
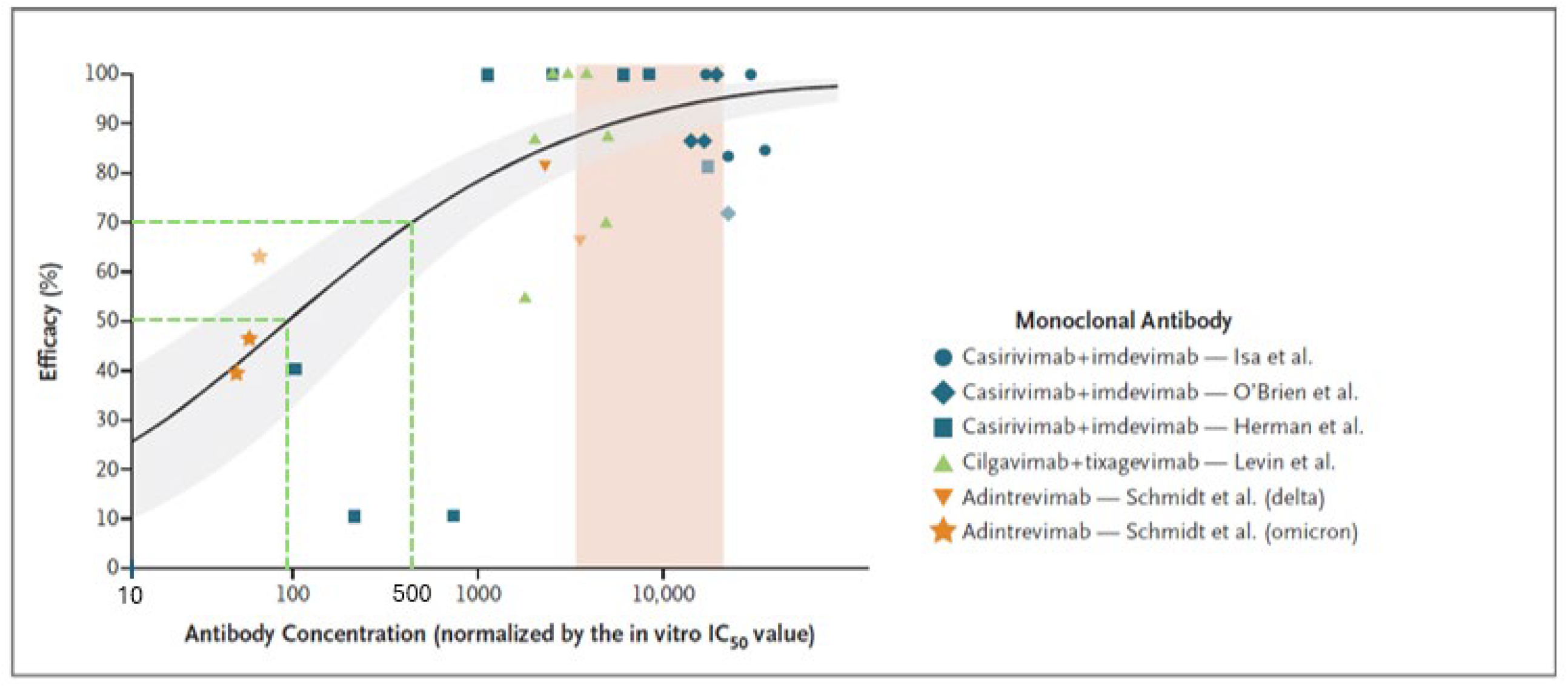
Neutralizing Titer Versus Efficacy Curve for relevant mABs. The black line represents estimated dose–response relationship determined by fitting a logistic model with maximal efficacy of 100% to the data, and the gray shaded area represents 95% confidence interval estimated with parametric bootstrapping. The vertical beige shaded area shows the range of neutralizing titers after IV administration of a single dose of 4500 mg pemivibart at the end of infusion on day 1, at month 1, and at month 3. The green dashed lines highlight antibody concentrations corresponding to efficacy levels of 50% and 70%.

It is important to note that, according to the Cox model, as titers increase above 1:500, the corresponding gains in efficacy diminish. For instance, doubling the titer from 1:50 to 1:100 results in a 6% increase in relative efficacy in non-IC, while increasing the titer from 1:500 to 1:1000 only yields a 3% increase (Table 2). A similar trend is observed in Figure 3, where efficacy shows diminishing effect above titers of approximately 1:500, suggesting that further increases may offer limited additional benefit. Therefore, both methods indicate that very high titers, which would require significantly higher mAb doses and more burdensome route of administration (i.e., IV), may not be preferable from a risk-benefit and healthcare utilization perspective.

The lower efficacy observed in Cohort A (IC) in comparison to the Cohort B (non-IC) (Figure 2) align with results from the prevalence-adjusted titer-based threshold of protection (ToP) model developed using data from the PROVENT (evaluation of tixagevimab/cilgavimab in non-IC participants) and SUPERNOVA (evaluation of sipavibart in IC participants) studies (Edge 2025). In that model, a titer of approximately 1:200 corresponds to 40% efficacy when IC and non-IC data were analyzed together; analysis of IC only was not performed. The CANOPY Cohort A model predicts a titer of 1:200 associated with 45% efficacy for IC population (Table 2), which is similar to the ToP model. Models developed using data from one platform (e.g., AstraZeneca platform) may have limitations in applicability to other mAbs (e.g., Invivyd platform) given differences in Fc modifications, innate effector functions, specific epitopes targeted, and other biophysical properties inherent to mAbs. Nevertheless, data such as these provide useful information to further support the novel data analytics employed with the CANOPY dataset to ascertain a reasonable titer correlate of protection.

Based on the Cox model, a titer target of approximately 1:500 for dominant circulating variants with the highest IC_50_ would ensure coverage against the spectrum of relevant circulating variants representing a range of IC_50_ values, as well as confer clinically meaningful protection in both IC and non-IC patients. Moreover, this target also safeguards against variant evolution, where a reduction in titer by approximately half (i.e., 1:250) equates to less than a 5% decrease in efficacy for the IC and non-IC populations. As higher titer targets (e.g., >1:500) necessitate higher overall administered dose with limited efficacy gain based on the Cox model, a titer target of 1:500 is reasonable and supported from both an efficacy and risk-benefit perspective in both IC and non-IC populations.

Following selection of a titer target, immunobridging calculations can be employed. For example, CANOPY utilized the lower limit of the 90% CI of the GMR between the selected titer target and the mAb/variant-specific neutralizing titer >0.8.

Importantly, as the Cox model is derived from the pemivibart efficacy data through 12 months and is variant-agnostic based on implementation of a weighted IC_50_, it can serve as a predictive tool for establishing platform-relevant (i.e., mAbs engineered with same structural backbone, Fc region modifications, and associated effector functions) titer targets and corresponding doses with which to immunobridge any future Invivyd platform mAbs with a high degree of certainty of translated clinical benefit.

## Conclusion

The data from CANOPY provide a robust foundation, incorporating both neutralizing titers and long-term clinical efficacy, to support a re-evaluation of the target titers used to guide dosing, predict estimated clinical benefit, and support immunobridging. This surrogate approach can be effectively derived using the proposed Cox model.

## Data Availability

All data produced in the present study are available upon reasonable request to the authors

## Ethics and Reporting

The authors declare that all relevant ethical guidelines have been followed, all necessary IRB and/or ethics committee approvals have been obtained, all necessary patient/participant consent has been obtained and the appropriate institutional forms archived for referenced trials supported by Invivyd, Inc.

